# Implementation of an Integrated Cardiology/Pharmacy GLP-1 Receptor Antagonist Prescribing to Improve Utilization and Cardiovascular Risk

**DOI:** 10.1101/2025.08.01.25332846

**Authors:** Daniel L. Pollmann, Jayne Parry, Ellen Cravero, Gretchen Benson, Otto A. Sanchez, Susan White, Larissa Stanberry, Carolyn Wambach, Michael D. Miedema

## Abstract

**Background:** Glucagon-like peptide-1 receptor agonists (GLP-1 RAs) are effective therapies for cardiovascular risk reduction, including in non-diabetic populations. However, utilization is hindered by high costs, poor insurance coverage, supply issues, and complex dosing regimens.

**Methods:** This single-center, retrospective cohort study included 385 eligible patients referred to a multidisciplinary cardiology/pharmacy GLP-1 RA program between June 2023 and March 2024. Pharmacists selected medications, managed insurance processes, and conducted follow-up for titration and adherence. Primary endpoints were rates of initiation and continuation. Secondary outcomes were changes in cardiovascular risk factors.

**Results:** Two thirds of patients referred to the program initiated GLP-1 RA therapy, and over 80% who followed up continued their medication, experiencing statistically significant reductions in weight, systolic blood pressure, and hemoglobin A1C.

**Conclusions:** This integrated cardiology/pharmacy GLP-1 RA program saw high rates of medication initiation and continuation and was associated with improvements in cardiovascular risk factors. Incorporating pharmacists into cardiovascular care can overcome key prescribing barriers and improve utilization of GLP-1 RAs to improve cardiovascular risk.

**WHAT IS KNOWN:**

- GLP-1 receptor agonists are underutilized in cardiovascular care despite compelling evidence for risk reduction.
- Barriers such as insurance denials, prior authorizations, high out-of-pocket costs, and complex titration regimens contribute to poor real-world initiation and persistence.

**WHAT THE STUDY ADDS:**

- Integrating pharmacists into cardiology prescribing workflows can overcome these barriers and significantly improve GLP-1 RA utilization.
- Improved access and persistence with GLP-1 RAs may help translate clinical trial evidence into real-world cardiovascular risk reduction.
- This model demonstrates a scalable approach to improving access to cardiometabolic therapies in high-risk populations.

## INTRODUCTION

Glucagon-like peptide-1 receptor agonists (GLP-1 RAs) have been shown to improve glycemic control, stimulate weight loss, and lead to a reduction in atherosclerotic cardiovascular disease (ASCVD) events, including in individuals without diabetes.^1,2^ Despite their proven clinical efficacy, several logistical and clinical challenges may prevent successful prescription and administration of GLP-1 RAs, as described by data from the American Diabetes Association and Academy of Managed Care Pharmacy.^3,4^ Their elevated cost leads to poor formulary coverage, a high burden of required prior authorizations, and frequent denials by payers. High demand has led to periodic supply issues leading to inconsistent use of certain formulations. Also, they require multiple dose titrations during a ramp-up period and can produce gastrointestinal (GI) side effects requiring additional dose adjustments, especially early in treatment. These barriers have led to prescriber hesitation and real-world adherence and persistence rates of GLP-1 RAs far below those reported in clinical trials.

Similar challenges for other medication classes and disease states with high cost and complex management requirements have been effectively addressed by involving pharmacists in prescription and monitoring workflows, leading to improved patient outcomes.^5–7^ For this analysis, we report the results of an integrated cardiology /pharmacy GLP-1 RA program to assess rates of initiation and continuation of prescribed GLP-1 RAs.

## METHODS

This was a single-center retrospective cohort study analyzing patients referred to the Allina Health Minneapolis Heart Institute (MHI) integrated cardiology/pharmacy GLP-1 RA program. In June of 2023, a multidisciplinary program involving staff cardiologists, advanced practice providers, and a pharmacy team at MHI clinics was devised to address challenges and improve efficiency in prescribing GLP-1 RAs. Patients were eligible for referral to the integrated cardiology/pharmacy GLP-1 RA program if they met the following eligibility criteria: body mass index (BMI) greater than 30 kg/m^2^ and established clinical cardiovascular disease (coronary artery disease, peripheral arterial disease, or cerebrovascular disease), type 2 diabetes, or multiple cardiovascular risk factors. Following patient referral by a cardiologist or cardiovascular advanced practice provider, the pharmacist at Allina Health MHI initiated the next steps starting with confirming eligibility criteria. Patients were then contacted by the pharmacist, and an appropriate GLP-1 RA was prescribed based on drug availability, insurance formularies, health history including diabetes and obesity status, cost, and patient preference. The pharmacist completed any required correspondence with insurance, including prior authorizations or appeal letters. Following successful initiation of the GLP-1 RA, the pharmacist called each patient every two to three weeks to review adherence and side effects. Dose titrations were made as necessary based on the medication schedule or side effects.

The primary endpoints were the rate of initiation of GLP-1 RAs and the rate of continuation among initiated patients with success prespecified at 50 and 75 percent, respectively. Reasons for failure of initiation or continuation of medications and all reported adverse effects were recorded. Secondary endpoints were changes in weight, blood pressure, lipid profile, and hemoglobin A1c from baseline. Blood pressure and laboratory values were recorded from clinic visits. Weight was also recorded at clinic visits and self-reported during phone interviews. Following initial data collection, a minimum follow-up period was set at one month for the primary endpoint of medication continuation and all secondary endpoints. Data was collected from electronic medical records of patients referred to the program who consented to inclusion in the study. All patient information was stored confidentially in a REDCap database. Study design and implementation were subject to approval and oversight by the local Institutional Review Board.

### Statistical methods

Descriptive variables were associated with percentages of totals. Values were recorded as median (IQ range) to lessen the effect of skewing from outliers. P-values comparing patient demographics and clinical characteristics between patients initiated versus not initiated on GLP-1 RAs were calculated using the Wilcoxon rank sum test, Pearson’s Chi-squared test, and Fisher’s exact test, where appropriate. P-values comparing secondary outcome variables to baseline were calculated using the paired t-test.

## RESULTS

### Sample size and demographics

Between June of 2023 and March of 2024, 385 eligible patients were referred to the integrated cardiology/pharmacy GLP-1 RA program, and 259 (67%) were successfully initiated on a GLP-1 RA. The demographics and baseline characteristics of all patients referred to the program, those initiated on a GLP-1 RA, and those who did not initiate a GLP-1 RA are shown in Table 1. Patients had a median age of 62 years (interquartile (IQ) range, 52-69 years), 56% were male, 41% had diabetes, and the median BMI was 39 (IQ range 35, 44). *Medication initiation and continuation*

**Table 1.** Demographics and clinical characteristics of patients referred to the integrated Cardiology/Pharmacy GLP-1 RA program. Values are expressed as median (IQ range). P-values were calculated using the Wilcoxon rank sum test, Pearson’s Chi-squared test, and Fisher’s exact test, where appropriate.

| CHARACTERISTIC | REFERRED<br>N = 385<sup>1</sup> | DID NOT INITIATE<br>N = 126<sup>1</sup> | INITIATED<br>N = 259<sup>1</sup> | P-VALUE |
| --- | --- | --- | --- | --- |
| Age | 62 (52, 69) | 63 (53, 70) | 61 (52, 68) | 0.3 |
| male | 217 (56%) | 76 (60%) | 141 (54%) | 0.3 |
| Race |  |  |  | >0.9 |
| Asian | 2 (0.5%) | 0 (0%) | 2 (0.8%) |  |
| Black | 18 (4.7%) | 5 (4.0%) | 13 (5.0%) |  |
| Native American | 4 (1.0%) | 1 (0.8%) | 3 (1.2%) |  |
| White | 359 (94%) | 119 (95%) | 240 (93%) |  |
| Height (m) | 1.73 (1.65, 1.80) | 1.75 (1.65, 1.83) | 1.73 (1.65, 1.80) | 0.7 |
| Weight (kg) | 118 (102, 135) | 120 (101, 133) | 117 (102, 137) | 0.6 |
| BMI (kg/m<sup>2</sup>) | 39 (35, 44) | 38 (35, 44) | 39 (34, 45) | 0.6 |
| SBP (mmHg) | 126 (118, 140) | 127 (120, 142) | 126 (115, 138) | 0.2 |
| DBP (mmHg) | 78 (70, 84) | 78 (70, 86) | 78 (70, 82) | 0.2 |
| Tobacco use status |  |  |  | 0.6 |
| Current | 29 (7.6%) | 7 (5.8%) | 22 (8.5%) |  |
| Former | 159 (42%) | 50 (41%) | 109 (42%) |  |
| Never | 192 (51%) | 64 (53%) | 128 (49%) |  |
| Diabetes status |  |  |  | <0.001 |
| No | 132 (34%) | 58 (47%) | 74 (29%) |  |
| Prediabetes | 93 (24%) | 30 (24%) | 63 (24%) |  |
| Yes | 158 (41%) | 36 (29%) | 122 (47%) |  |

Initiation of GLP-1 RA therapy did vary by diabetes status, with 77% (122 out of 158) of individuals with diabetes initiated on a GLP-1 RA compared to 61% (137 out of 225) in patient without diabetes (p <0.01). Among the patients initiated on a GLP-1 RA with minimum one month follow-up, 198 (84%) continued therapy with a median duration of follow-up of 154 days (∼ 5 months) by the end of the data collection period (IQ range 98, 203). Of the 37 (16%) who did not, the median duration on a GLP-1 RA was 146 days (IQ range 93, 173). Reasons for discontinuation were lack of insurance coverage (12), medication intolerance (10), recent or anticipated change in clinical status (7), cost (3), medication unavailability (2), and other reasons (3) (Figure 1). Of the 126 patients (33%) who did not initiate GLP-1 RA therapy, the most common reasons were lack of payer coverage (53), cost to patient (21), patient disinterested or declined (17), inability to reach patient (14), patient hesitancy regarding injectable medications or potential side effects (9), and medication unavailability (6). A flowchart summary of initiation and continuation outcomes is found in Figure 1. The GLP-1 RA most initiated among patients was Ozempic (semaglutide, 46%) followed by Zepbound (tirzepatide, 22%), Wegovy (semaglutide, 19%), and all others (13%).

**Figure 1.**
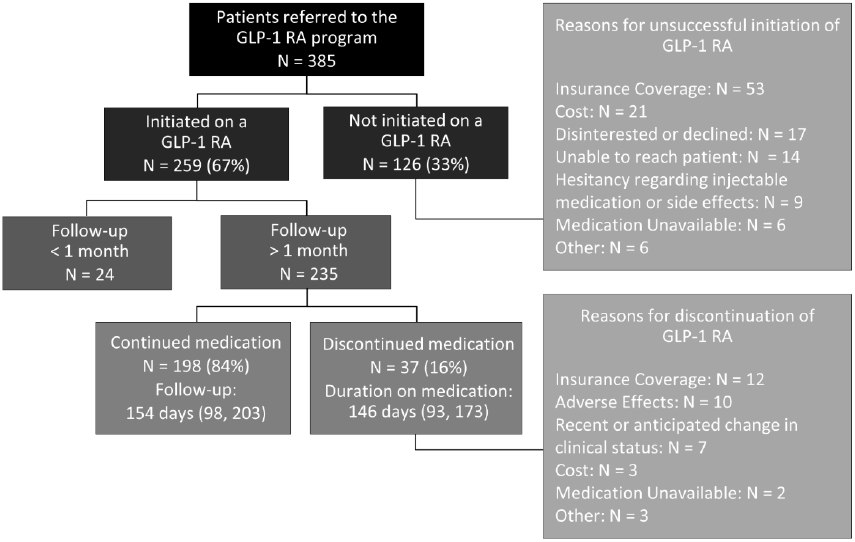
Initiation and continuation outcomes for the integrated cardiology/pharmacy GLP-1 RA program. Values are expressed as median (IQ range).

### Impact on weight and other CVD risk factors

Of 259 patients who initiated a GLP-1 RA, 235 (91%) had a minimum one-month follow-up. The impacts of GLP-1 RA therapy on cardiovascular risk factors in these patients are shown in Table 2. The median change in weight was –8 kg (–7% of starting body weight), corresponding to a median change in BMI of –2.86 kg/m^2^. Systolic and diastolic blood pressure changed by a median of –7 and –2 mmHg, respectively. Total cholesterol and triglycerides fell by a median of 16 and 13 mg/dL (10% and 9%), respectively, and hemoglobin A1C fell by a median of 0.45%.

**Table 2.** Secondary endpoints for patients successfully initiated and continued GLP-1 RAs with minimum one month follow-up. Values are expressed as median (IQ range). P-values were calculated using the paired t-test.

| CHARACTERISTIC | N = 198 | P-VALUES |
| --- | --- | --- |
| Weight, n (%) | 198 (100) |  |
| Time to follow-up, days | 155 (100, 211) |  |
| Change in weight, kg | -8 (-13, -5) | <0.001 |
| Change in BMI, kg/m<sup>2</sup> | -2.9 (-4.4, -1.7) | 0.39 |
| Blood glucose, n (%) | 134 (68) |  |
| Time to follow-up, days | 119 (69, 191) |  |
| Change in blood glucose, mg/dL | -13 (-29, -4) | <0.001 |
| Hemoglobin A1C, n (%) | 112 (57) |  |
| Time to follow-up, days | 114 (75, 176) |  |
| Change in hemoglobin A1C, % | -0.45 (-0.80, -0.18) | <0.001 |
| Blood pressure, n (%) | 150 (76) |  |
| Time to follow-up, days | 142 (97, 200) |  |
| Change in SBP, mmHg | -7 (-16, 6) | 0.003 |
| Change in DBP, mmHg | -2 (-9, 4) | 0.063 |
| Lipids, n (%) | 117 (59) |  |
| Time to follow-up, days | 119 (65, 177) |  |
| Change in total cholesterol, mg/dL | -16 (-33, 4) | <0.001 |
| Change in triglycerides, mg/dL | -13 (-65, 16) | <0.001 |

### Side effects

The most frequently reported side effects from those taking GLP-1 RAs (Figure 2) were nausea (33%), diarrhea (20%), and constipation (19%). The team pharmacist responded to persistent and intolerable side effects with dose adjustment recommendations; 10 patients (5%) discontinued their medications due to side effects.

**Figure 2.**
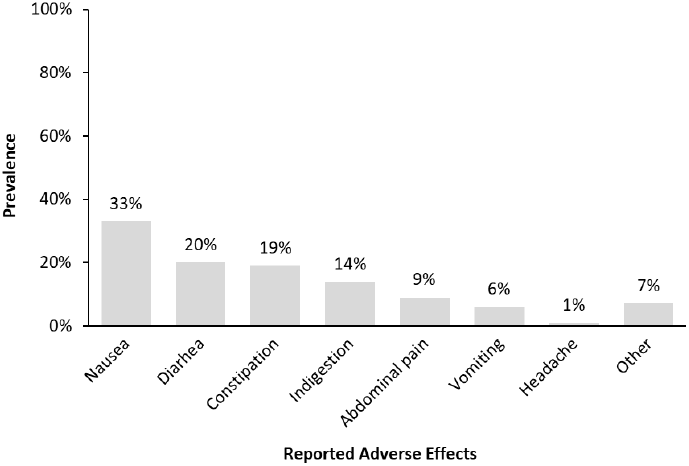
Prevalence of reported adverse effects among patients initiated on GLP-1 RAs with minimum one month follow-up (N = 235).

## DISCUSSION

This study analyzed a single-center, integrated cardiology/pharmacy GLP-1 RA program, a novel multidisciplinary approach designed to address prescribing challenges, improve access for patients with and without diabetes, and ultimately enhance cardiovascular health. The program resulted in a high rate of initiation, with over 2/3 of patients starting a GLP-1 RA. More than 80% of participants continued the medication during the follow-up period resulting in significant reductions in weight and several other CVD risk factors. Integration of pharmacy-based programs for GLP-1 RA medications into routine cardiovascular care can increase optimal utilization of these therapies and improve cardiovascular health.

Prior efforts to optimize GLP-1 RA utilization by leveraging pharmacist support have focused solely on patients with diabetes or elevated BMI,^6,8–10^ and have reported initiation rates varying from 19% to 51%. Notably, the highest reported percent success rate was based on pooled prescriptions of both GLP-1 RAs and SGLT2 inhibitors.^9^ Our program was evaluated during a period of significant GLP-1 RA shortages due to surging demand and global supply chain disruptions partly related to the COVID-19 pandemic.^11^ Despite these and other challenges, including payer denials, high costs, and prior authorizations, the assistance from a clinical pharmacist led to a high rate of initiation of a GLP-1 RA therapy, including in those without diabetes.

Additionally, despite the program occurring during the aforementioned challenges of medication shortages and declining insurance coverage, our analysis found high rates of continuation (>80%). Recent data from a large cohort of US healthcare systems found that the continuation rate at one year for GLP-1 RAs in patients without diabetes was only 35%, while patients with diabetes had a better, but still suboptimal, continuation rate of 54%.^12^ Continuation rates at six months in other studies have varied from 15% to 75%.^6,13–15^ There is a complex array of barriers to GLP-1 RA continuation, including high cost, adverse effects, aversion to injection, frequent dosing, and titration schedules. However, low starting doses and simplified administration regimens are linked to improved rates of continuation.^13^ Pharmacists are well-equipped to advise patients on optimal GLP-1 RA selection and optimal methods for titration. The ongoing contact from pharmacists helps patients navigate difficulties in the titration and maintenance phases of their therapy, tasks that if left to the provider, may lead to hesitancy in prescribing GLP-1 RAs.

For the 33% of patients not initiated on a medication, the two most common reasons – lack of insurance coverage and high patient costs – were typical of findings in other studies of GLP-1 RA prescribing trends.^6,8,9^ Moreover, the significant difference in diabetes status between patients who were initiated versus not initiated on GLP-1 RAs also aligns with the data, likely due to the relative ease of prescribing diabetes formulations of this medication class compared to alternatives. For the 16% who discontinued GLP-1 RA therapy, the reasons were consistent with prior findings and reflect the known challenges associated with this medication class, including insurance coverage and adverse effects.^6,13–15^ The adverse effects observed here were similar to those reported in the large randomized trials for several of these agents.^1,16,17^

In our program, cardiovascular risk factors were reduced as expected based on randomized controlled data of this medication class in patients with and without diabetes.^1,2,16–18^ This suggests that there is real-world ASCVD risk reduction for patients who are successfully continued these medications.

## LIMITATIONS

A primary limitation of this study is its retrospective observational design and the absence of a controlled comparison group, which reduces confidence in the superiority of this program over conventional prescribing methods. However, the rates of initiation and continuation were significantly higher than those reported in other studies.

Our average follow-up time (∼5 months) was shorter than that of other analyses, and the rate of discontinuation may be higher at 1 year. However, a prior real-world retrospective study of GLP-1 RA prescriptions in the U.S. indicated that the steepest drop-off in persistence occurred in the first month.^14^

Our sample size was modest but adequate for a hypothesis-generating study that supports the implementation of larger, integrated programs between pharmacy, cardiology, and primary care to optimize the use of GLP-1 RAs. Finally, self-reporting of body weights by patients during telehealth appointments or follow-up calls with the pharmacist introduces potential opportunities for measurement error or false reporting. Given the modest weight loss observed in our cohort compared to others,^6^ it is unlikely that this effect was exaggerated in our study.

## CONCLUSIONS

In conclusion, the implementation of an integrated program between cardiology and pharmacy to optimize the use of GLP-1 RAs in patients with appropriate clinical indications resulted in rates of initiation and continuation that were substantially higher than those reported in the general population. Health care systems should evaluate how best to optimize delivery programs for GLP-1 RA’s given their significant impact on cardiovascular health.

## Data Availability

Data will be available within the article or viewable by request in an appropriate repository at time of publication.

## ACKNOWLEDGEMENTS

The authors would like to thank the cardiology providers, advanced practice clinicians, and pharmacy team at Allina Health Minneapolis Heart Institute for their collaboration in designing and implementing the integrated GLP-1 RA program. We also acknowledge the support of the Minneapolis Heart Institute Foundation and data analytics team for assistance with data collection and management. We are grateful to the patients who participated in this program and contributed to the evaluation of its outcomes.

## NONSTANDARD ABBREVIATIONS AND ACRONYMS

GLP-1 RA: Glucagon-like peptide-1 receptor agonist
ASCVD: Atherosclerotic cardiovascular disease
MHI: Allina Health Minneapolis Heart Institute
IQ: Interquartile
PCSK9: proprotein convertase subtilisin/kexin type 9
SGLT2: Sodium-Glucose Cotransporter 2.

